# The evidence supporting AHA guidelines on adult cardiopulmonary resuscitation (CPR)

**DOI:** 10.1101/2023.12.07.23299702

**Authors:** Emma Junedahl, Peter Lundgren, Erik Andersson, Vibha Gupta, Truls Råmunddal, Aidin Rawshani, Gabriel Riva, Ida Arnetorp, Fredrik Hessulf, Johan Herlitz, Therese Djärv, Araz Rawshani

## Abstract

**Background:** Guidelines for the management of cardiac arrest play a crucial role in guiding clinical decisions and care. We examined the strength and quality of evidence underlying these recommendations in order to elucidate strengths and gaps in knowledge.

**Methods:** Using the 2020 American Heart Association (AHA) Guidelines for Adult CPR, we subdivided all recommendations into advanced life support (ALS), basic life support (BLS), and recovery after cardiac arrest, as well as a more granular categorization by topic (i.e. the intervention or evaluation recommended). The Class of Recommendation (COR) and Level of Evidence (LOE) for each were reviewed.

**Results:** We noted 254 recommendations, of which 181 were ALS, 69 were BLS, and 4 were recovery after resuscitation. In total, only 2 (0.8%) had the most robust evidence (LOE A), while 23% were at LOE B-NR(Non-Randomized), 15% at LOE B-R (Randomized), 50% at LOE C-LD (Limited Data), and 12% relied on expert opinion LOE C-EO (Expert Opinion). Despite the strength of ALS recommendations (Class 1, 2a, or 2b), none had LOE A. In BLS, no recommendations were supported by LOE A. For BLS, 74% of recommendations had LOE C (C-LD or C-EO). The evidence for specific BLS topics, such as airway management, was notably low. Among ALS topics, neurological prognostication had relatively stronger evidence. Overall, 26.1% of BLS recommendations had LOE A or B, versus 43.1% for ALS recommendations.

**Conclusions:** There is a strong discrepancy between the strength of recommendation and the underlying evidence in cardiac arrest guidelines. The findings underscore a pressing need for more rigorous research, particularly randomized trials.

## Introduction

Cardiac arrest impacts 400,000 Europeans and 350,000 Americans annually, of which 90% succumb.^1^ The International Liaison Committee on Resuscitation (ILCOR) issues evidence-based treatment guidelines for all aspects of cardiopulmonary resuscitation. Every five years, ILCOR releases updated guidelines on cardiopulmonary resuscitation (CPR) and emergency cardiovascular care to mirror the most recent scientific advances.^2^ The European Resuscitation Council (ERC) and American Heart Association (AHA) use the ILCOR statements to develop clinical practice guidelines.

Recommendations are divided into different classes of recommendation (COR) and levels of evidence (LOE). The COR rating reflects the benefit-risk balance and corresponds to the strength of the recommendation. Class 1 indicates that an assessment or intervention is useful, effective, and should be used. Class 2 recommendations are weaker and have a lower degree of benefit, compared to risk. For Class 2a, the benefit is estimated to marginally exceed the risk. Class 2b recommendations should be used selectively and individually, as the benefit is questionable. Class 3 implies that the potential benefit is as great as the risk, which suggests the absence of evidence or risk of harm.

Level A (LOE-A) involves high-quality evidence from multiple randomized controlled trials (RCT), meta-analyses of high-quality RCTs, and one or more RCTs supported by high-quality registry studies. Level B-Randomized (LOE B-R) is based on moderately qualitative evidence from one or more RCTs or meta-analyses of moderate-qualitative RCTs. Level B-Non Randomized (LOE B-NR) entails moderate qualitative evidence from one or more well-designed and well-performed non-randomized studies. Level C-Limited Data (LOE C-LD) is based on limited data and includes either randomized or non-randomized observational or registry studies with limitations in design or execution. Level C-Expert Opinion (LOE C-EO) is based on expert opinion.^3^

A review published in 2009 of the ACC/AHA guidelines between 1984-2008 showed that 11% of the recommendations were classified as LOE A.^4^ A review of the recommendations issued between 2008-2018, showed that the corresponding proportion was 8.5%.^5^ We conducted a renewed survey of the latest guidelines, published in 2020 to map the development of the evidence as well as variations in different aspects of CPR in adults.

## Methods

Data collected from the *American Heart Association* (AHA) *Guidelines for CPR and Emergency Cardiovascular Care* 2020 Part 3 Adult CPR.^3^

All individual recommendations issued in 2020 were assigned a category relating to the topic, of which there were the following: Accidental hypothermia, Adjuncts to CPR, Airway management, Amiodoarone or lidocaine, Aanaphylaxis or asthma, Antiarrhythmic agents excluding amiodarone and lidocaine, BLS recognition and initiation of CPR, Cardiac arrest due to overdose and toxicity, Cardiac arrest in cardiac surgery, Cardiac arrest in special circumstances, Chest compressions, Coronary angiography, CPR feedback, monitoring, checks, Defibrillation, Drowning, ECMO (extracorporeal membrane oxygenation), Electrolyte abnormalities in cardiac arrest, Glycemic control post-ROSC, Hemodynamic management, post-ROSC, Initiation of CPR, Management of bradycardia, Management of supra ventricular arrythmias, Neuroprognostication, Other, Other antiarrythmic interventions, Other pharmacological agents, Positioning during CPR, Post ROSC diagnostics, Pregnancy, Prophylactic antibiotics post-ROSC, Pulmonary embolism, Recovery and survivorship, Seizure management, Steroids, Termination of resuscitation, Targeted temperature management (TTM), Vascular access, Vasopressors, Ventilation, Ventilation post-ROSC.

Additionally, the recommendations were subdivided into ALS (advanced life support), BLS (basic life support) and recovery. The COR and LOE were reviewed in each category to explore the strength of the recommendation and the underlying evidence. We also considered recommendations issued in 2021 and 2022 to determine whether previous recommendations (2020) had changed the level of evidence.

## Results

Among 254 recommendations, there were 181 recommendations for ALS, 69 recommendations for BLS, and 4 recommendations for Recovery after cardiac arrest.

Overall recommendations had the following distribution: 0.8% (2) had LOE A, 15% (37) had LOE B-R, 23% (58) had LOE B-NR, 50% (126) had LOE C-LD, and 12% (31) had LOE C-EO.

ALS recommendations had the following distribution: 1.1% (2) had LOE A, 17% (30) had LOE B-R, 25% (46) had LOE B-NR, 46% (84) had LOE C-LD, and 10% (19) had LOE C-EO.

BLS recommendations had the following distribution: 0% (0) had LOE A, 10% (7) had LOE B-R, 16% (11) had LOE B-NR, 57% (39) had LOE C-LD, and 17% (12) had LOE C-EO.

There were in total 32% (81) Class 1 recommendations. Among those, 0% (0) had LOE A, 2.8% (7) had LOE B-R, 7.5% (19) had LOE B-NR, 15% (39) had LOE C-LD, and 6.3% (16) had LOE C-EO.

There were in total 23% (58) Class 2a recommendations. Among those, 0% (0) had LOE A, 3.9% (10) had LOE B-R, 5.5% (14) had LOE B-NR, 12% (30) had LOE C-LD, and 1.6% (4) had LOE C-EO.

There were in total 35% (89) Class 2b recommendations. Among those, 0% (0) had LOE A, 5.1% (13) had LOE B-R, 7.5% (19) had LOE B-NR, 19% (48) had LOE C-LD, and 3.5% (9) had LOE C-EO.

There were in total 5.9% (15) Class 3: No benefit recommendations. Among those, 0.8% (2) had LOE A, 2.4% (6) had LOE B-R, 1.2% (3) had LOE B-NR, 1.6% (4) had LOE C-LD, and 0% (0) had LOE C-EO.

There were in total 4.3% (11) Class 3: Harm recommendations. Among those, 0% (0) had LOE A, 0.4% (1) had LOE B-R, 1.2% (3) had LOE B-NR, 2% (5) had LOE C-LD, and 0.8% (2) had LOE C-EO.

Figure 1a illustrates the distribution of class of recommendation and level of evidence for ALS, BLS, and recovery after cardiopulmonary resuscitation. We note that very few recommendations suggest harm or absence of benefit, whereas the majority suggest clear benefit (COR 1) or probable benefit (COR 2a). Yet, a negligible portion (0.8%) of these recommendations were based on firm evidence (LOE A). Overall, 26.1% of BLS recommendations had LOE A or B, as compared with 43.1% for ALS.

**Figure 1a.**
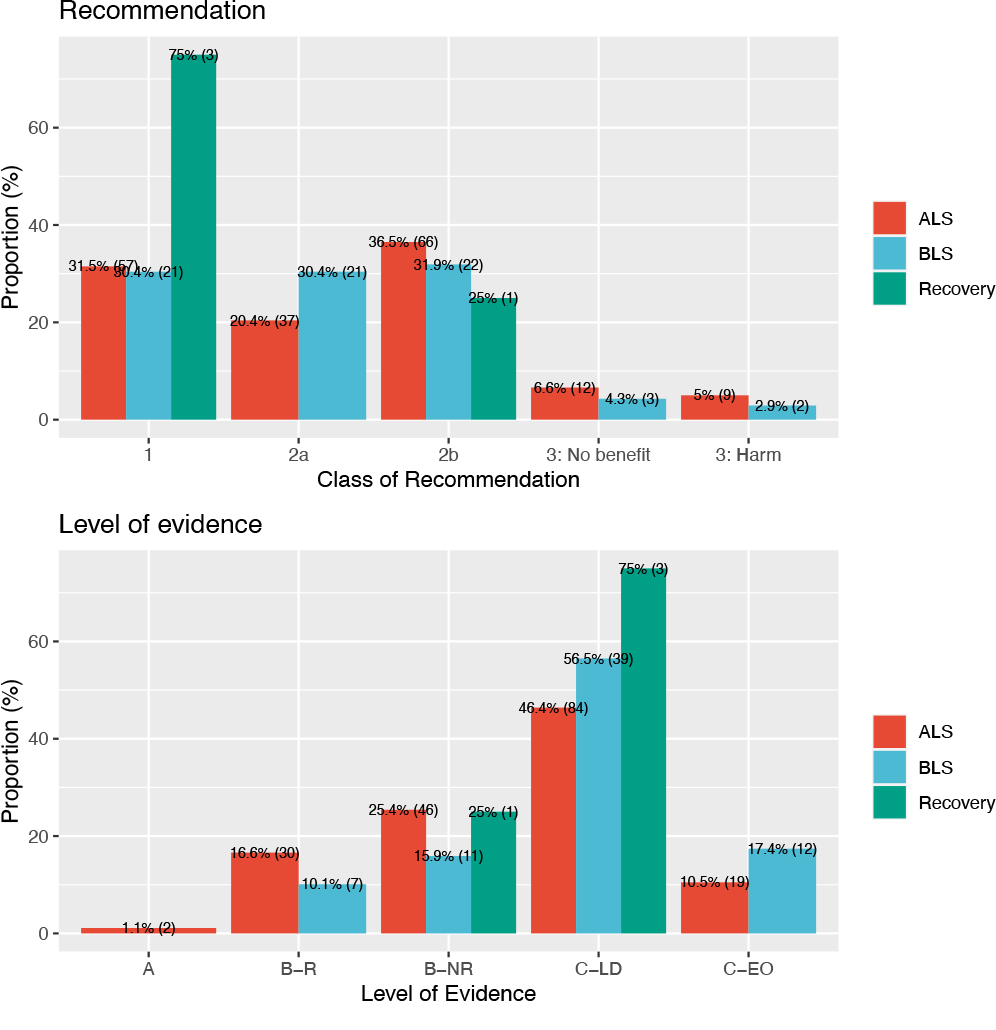
Distribution for recommendations and level of evidence across recommendations (ALS, BLS and recovery after CPR).

**Figure 1b.**
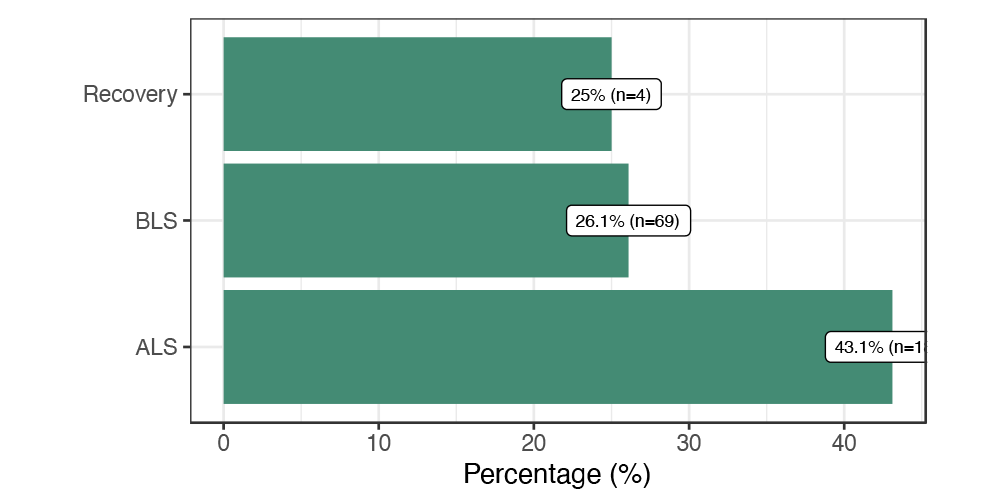
Proportion of recommendations that have LOE A or B.

Figure 2 displays the topic-specific level of evidence and class of recommendation. For BLS, all recommendations for airway management, positioning, and ventilation were based on low evidence (LOE C). Among BLS recommendations for defibrillation, 5 out of 16 recommendations had LOE B (B-NR or B-R). For ALS, neuroprognostication stood out as the most evidence-based topic, although all recommendations with LOE B were of type B-NR (i.e., the lower grade of LOE B).

**Figure 2.**
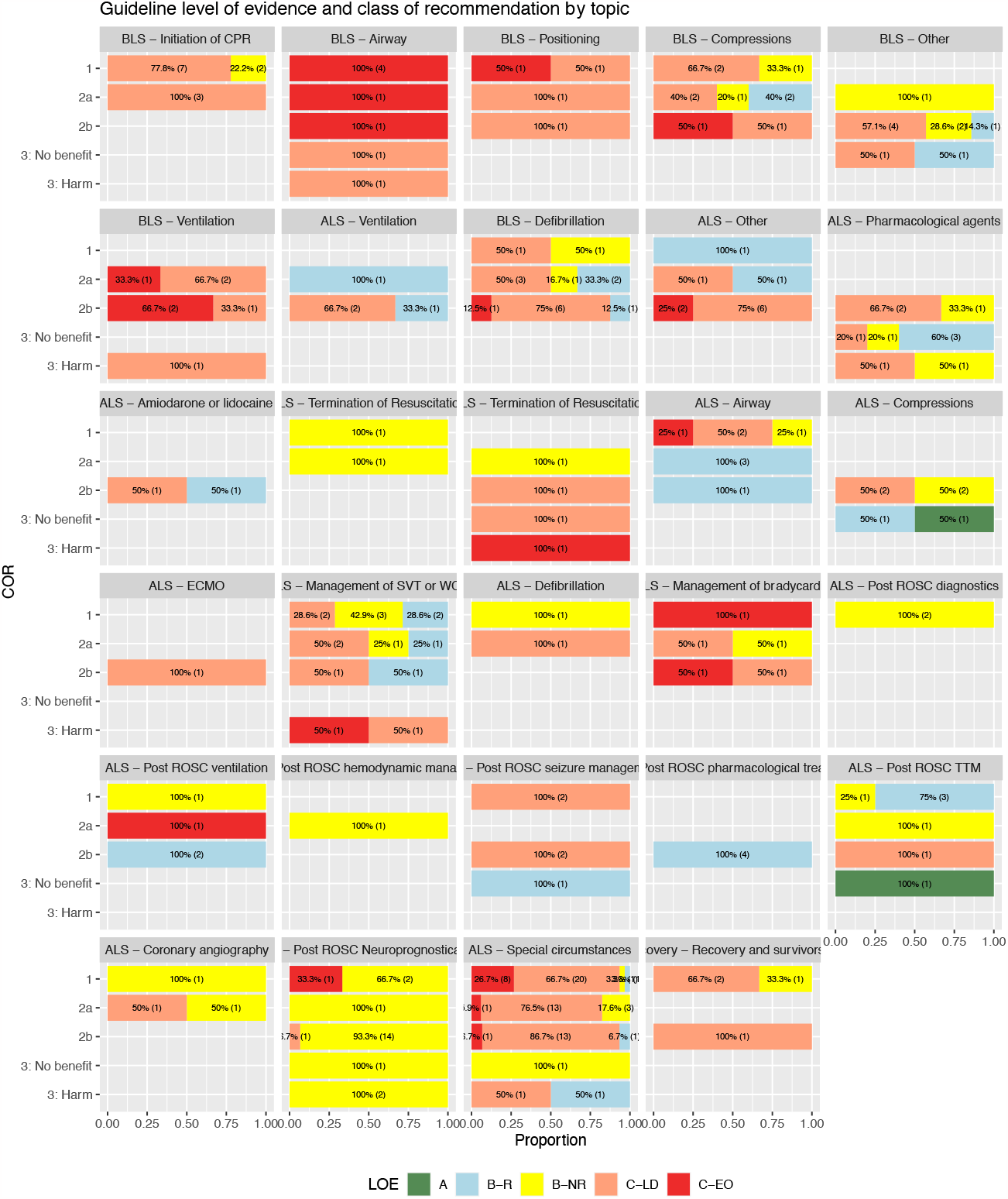

Figure 3 shows the proportion of all recommendations, by topic, which had at least LOE B (either B-R or B-NR). Recommendations for management post ROSC had a clearly higher evidence grade.

**Figure 3.**
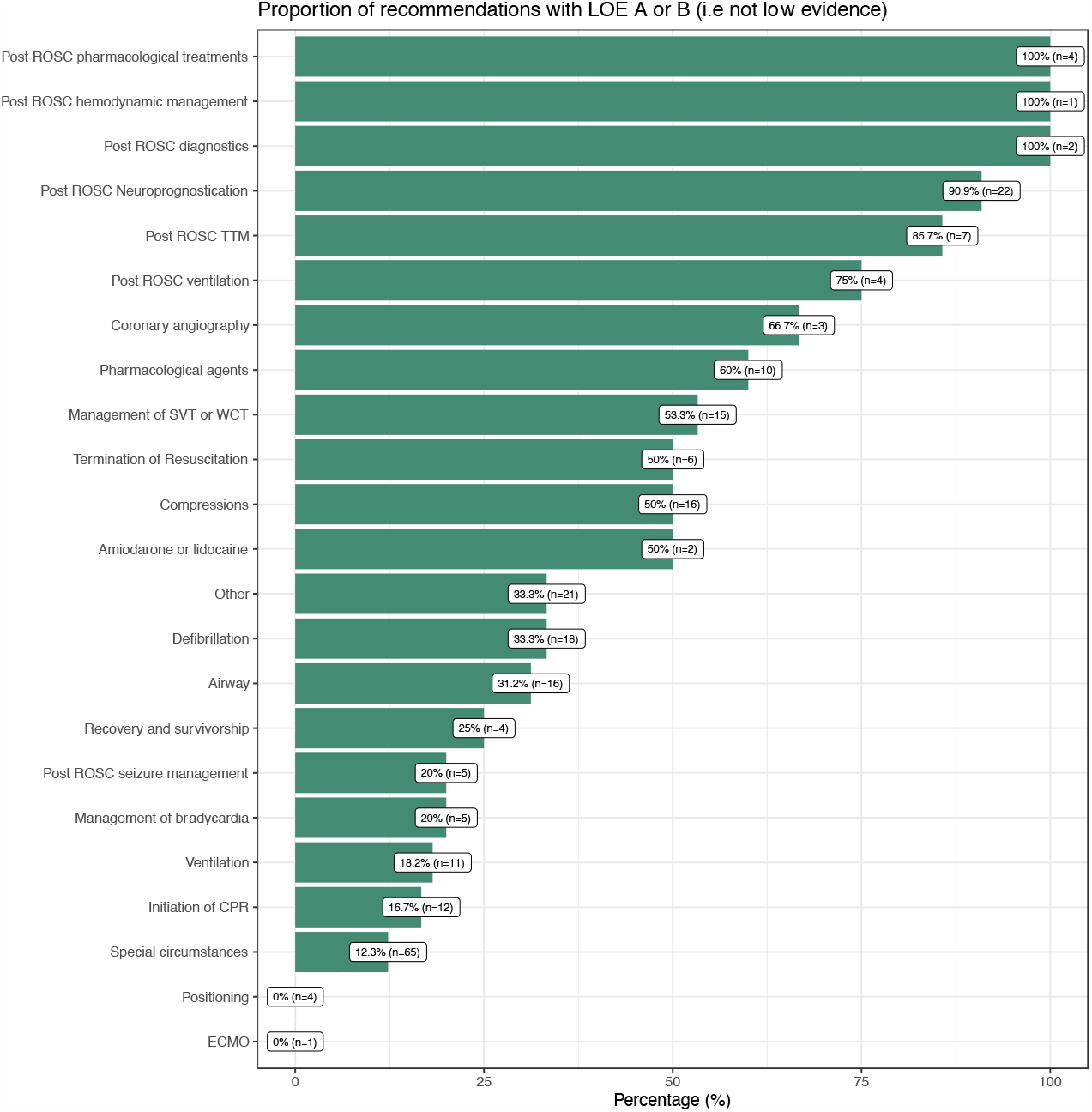
The number in parenthesis depicts the total number of recommendations available.

## Discussion

The evidence base for current guidelines on the management of cardiac arrest reveals a strong discrepancy between the strength class of recommendation and the underlying level of evidence. While a majority of recommendations suggest a clear or probable benefit (Class 1 and Class 2a), very few are backed by robust evidence. Such a mismatch underscores the challenges and gaps in producing high-quality evidence in the hyperacute, real-time scenarios of cardiac arrest; this is suggested by the difference in the proportion of recommendations with LOE A or B for the pre- and post-ROSC phase.

Among ALS recommendations, we note the absence of reliable evidence (LOE A) despite a high prevalence of strong recommendations (Class 1, 2a, or 2b). Over half of the ALS recommendations were based on limited data or expert opinions, which, while valuable and perhaps clinically sound, do not provide the rigorous evidence essential for such critical care settings. We recognize that lack of reliable evidence may have little clinical significance for some interventions (e.g. the recommendation to record a 12-lead ECG upon ROSC), but this study demonstrates an array of critical interventions and tasks that lack a sound evidence-base.

This simple study elucidates the significant gaps in our current understanding of cardiac arrest management, underscoring an urgent need for more studies focusing on these critical knowledge gaps. We emphasize that researchers should channel their efforts and resources towards more interventional studies, preferably in national and international collaborations to pool resources, expertise and power. We also recognize that while observational studies have their merits, there is a pressing need for more randomized trials and intervention studies.

With regards to BLS, perhaps the most widespread treatment ever introduced, there is an even more pronounced evidence deficit. With no recommendations supported by LOE A and a significant proportion relying on the weakest evidence types (LOE C-LD and LOE C-EO). We view this as a clear imperative to invest in more robust research within this domain, recognizing that there are many groups worldwide conducting excellent research in this field. Our study is in line with research in other cardiovascular conditions, as demonstrated by Tricoci et al,^4^ Fanaroff et al, who showed that only a minority of cardiovascular guidelines were based on LOE A.^5^ Althought, the Evolution of the ACC/AHA Clinical Guidelines study found that between 2008 and 2018, the ACC/AHA guidelines saw a decrease in LOE-C recommendations, suggesting that the guidelines were gradually strengthened in that mening that recommendations based on lower quality (LOE-C) were removed. However, no corresponding increase in higher level of evidence has yet to been seen.^6^

We also note relatively few Class 2b and Class 3 recommendations (lack of effect or potential harm) within the guidelines, which we believe is a reflection of the well-documented issue in scientific research: the under-reporting of negative or null results. Studies that fail to demonstrate significant beneficial outcomes or show potential adverse effects are often less likely to be submitted, accepted, or published compared to those with positive outcomes. We find no reason to believe the cardiac arrest field would differ from other fields regarding this, and it is presumably an important explanation for the distortion in the distribution demonstrated here. This gap not only affects the reliability of guidelines but also deprives clinicians and researchers of vital information that could guide clinical decision-making and future research.

In conclusion, the prevailing evidence base accentuates a pressing need for enhanced research efforts, with more focus on interventional trials. Both ALS and BLS recommendations, crucial in the hyperacute context of cardiac arrest, warrant more rigorous and high-quality evidence from randomized trials.

## Data Availability

To conduct this study, we utilized the American Heart Association (AHA) Guidelines for Adult CPR from 2020. All recommendations pertaining to advanced life support (ALS), basic life support (BLS), and recovery after cardiac arrest were reviewed according to the guidelines, and the classifications (Class of Recommendation, COR) and levels of evidence (Level of Evidence, LOE) for each recommendation were documented. This study is solely based on the analysis of and references to the publicly available 2020 AHA guidelines and does not involve separately collected data.

---

**Tables 1.** Level of evidence (LOE) and class of recommendation (COR) for all recommendations, ALS recommendations and BLS recommendations.

| All recommendations | Level of Evidence |  |  |  |  | Total |
| --- | --- | --- | --- | --- | --- | --- |
|  | A | B-R | B-NR | C-LD | C-EO |  |
| COR |  |  |  |  |  |  |
| 1 | 0 (0%) | 7 (2.8%) | 19 (7.5%) | 39 (15%) | 16 (6.3%) | 81 (32%) |
| 2a | 0 (0%) | 10 (3.9%) | 14 (5.5%) | 30 (12%) | 4 (1.6%) | 58 (23%) |
| 2b | 0 (0%) | 13 (5.1%) | 19 (7.5%) | 48 (19%) | 9 (3.5%) | 89 (35%) |
| 3: No benefit | 2 (0.8%) | 6 (2.4%) | 3 (1.2%) | 4 (1.6%) | 0 (0%) | 15 (5.9%) |
| 3: Harm | 0 (0%) | 1 (0.4%) | 3 (1.2%) | 5 (2.0%) | 2 (0.8%) | 11 (4.3%) |
| Total | 2 (0.8%) | 37 (15%) | 58 (23%) | 126 (50%) | 31 (12%) | 254 (100%) |

| ALS | Level of Evidence |  |  |  |  | Total |
| --- | --- | --- | --- | --- | --- | --- |
|  | A | B-R | B-NR | C-LD | C-EO |  |
| COR |  |  |  |  |  |  |
| 1 | 0 (0%) | 7 (3.9%) | 13 (7.2%) | 26 (14%) | 11 (6.1%) | 57 (31%) |
| 2a | 0 (0%) | 6 (3.3%) | 10 (5.5%) | 19 (10%) | 2 (1.1%) | 37 (20%) |
| 2b | 0 (0%) | 11 (6.1%) | 17 (9.4%) | 34 (19%) | 4 (2.2%) | 66 (36%) |
| 3: No benefit | 2 (1.1%) | 5 (2.8%) | 3 (1.7%) | 2 (1.1%) | 0 (0%) | 12 (6.6%) |
| 3: Harm | 0 (0%) | 1 (0.6%) | 3 (1.7%) | 3 (1.7%) | 2 (1.1%) | 9 (5.0%) |
| Total | 2 (1.1%) | 30 (17%) | 46 (25%) | 84 (46%) | 19 (10%) | 181 (100%) |

| BLS | Level of Evidence |  |  |  |  | Total |
| --- | --- | --- | --- | --- | --- | --- |
|  | A | B-R | B-NR | C-LD | C-EO |  |
| COR |  |  |  |  |  |  |
| 1 | 0 (0%) | 0 (0%) | 5 (7.2%) | 11 (16%) | 5 (7.2%) | 21 (30%) |
| 2a | 0 (0%) | 4 (5.8%) | 4 (5.8%) | 11 (16%) | 2 (2.9%) | 21 (30%) |
| 2b | 0 (0%) | 2 (2.9%) | 2 (2.9%) | 13 (19%) | 5 (7.2%) | 22 (32%) |
| 3: No benefit | 0 (0%) | 1 (1.4%) | 0 (0%) | 2 (2.9%) | 0 (0%) | 3 (4.3%) |
| 3: Harm | 0 (0%) | 0 (0%) | 0 (0%) | 2 (2.9%) | 0 (0%) | 2 (2.9%) |
| Total | 0 (0%) | 7 (10%) | 11 (16%) | 39 (57%) | 12 (17%) | 69 (100%) |

